# Association of COVID-19 risk factors with systemic fungal infections in hospitalized patients

**DOI:** 10.1101/2024.10.10.24315254

**Authors:** Abbygail C. Wilbourn, Oleg V. Tsodikov, Sylvie Garneau-Tsodikova

## Abstract

**Purpose:** A new category of systemic co-infections that emerged with the COVID-19 pandemic is known as COVID-19-associated (CA) fungal infections, which include pulmonary aspergillosis (CAPA), candidiasis (CAC), and mucormycosis (CAM). We aimed to study the association between patient characteristics of hospitalized COVID-19 patients, COVID-19 comorbidities, and COVID-19 therapies with secondary non-superficial fungal infections.

**Methods:** We performed descriptive and regression analyses of data from 4,999 hospitalized COVID-19 patients from the University of Kentucky Healthcare (UKHC) system.

**Results:** The patients with secondary systemic fungal infections had a 6-fold higher risk of death than those without such infections. Generally, the risk factors for severe COVID-19 (age, obesity, cardiovascular disease, diabetes, and lack of COVID-19 vaccination) were strong predictors of a secondary fungal infection. However, several characteristics had much higher risks, suggesting that a causative link may be at play: ICU admission, mechanical ventilation, length of hospital stay, and steroid use.

**Conclusions:** In sum, this study found that the known risk factors for severe COVID-19 disease, age, diabetes, cardiovascular disease, obesity, ventilation, and high steroid doses were all predictors of a secondary fungal infection. Steroid therapy may need to be modified to account for a risk or a presence of a fungal infection in vulnerable patients.

## Introduction

The severe acute respiratory syndrome coronavirus 2 (SARS-CoV-2) is a member of the same family as the previously identified severe acute respiratory syndrome (SARS) coronavirus and the Middle East respiratory syndrome (MERS) coronavirus. The SARS-CoV-2 virus, originally isolated in Wuhan, China in December of 2019, caused coronavirus disease known as COVID-19 [1]. COVID-19 spread very quickly throughout the world causing over 760 million confirmed cases and nearly 7 million deaths as of April 2023 [2]. Early on, COVID-19-associated pulmonary aspergillosis (CAPA), an invasive fungal infection, was reported [3]. High mortality rates ascribed to this comorbidity prompted several subsequent studies suggesting that corticosteroids used in COVID-19 therapy were an underlying contributor to fungal infections [4, 5]. Steroids were also noted as a risk factor for fungal infections linked to influenza-associated pneumonia [6]. Similarly, an increase in COVID-19-associated mucormycosis (CAM) in severely ill patients in the second wave of pandemic caught the attention of researchers, as a significant complication, leading to higher risk of death or disfigurement due to surgical procedures [7]. The incidence of invasive fungal infections among COVID-19 patients is especially high in the intensive care units (ICUs), where it is in the range of 5-20% as reported in different studies [8]. The inflammation and dysregulation of the immune system caused by COVID-19 are thought to increase the risks of fungal infections by creating favorable conditions for fungal invasion [9].

While most common COVID-19 symptoms are respiratory, this disease can affect the human body systemically, with a wide range of symptoms from respiratory to gastrointestinal. The severity of these symptoms differs from person to person based on comorbidities and the immune response to the virus. It is widely accepted that contracting SARS-CoV-2 causes dysregulation in the immune system and promotion of inflammatory thrombosis *via* the coagulation cascade [10]. The viral RNA causes dysregulation of interferons [11–14]. SARS-CoV-2 also dysregulates B and T cells [15, 16]. In addition to the effects of the virus itself that likely favor fungal infections, therapeutic interventions against COVID-19 can also contribute. During the beginning of the pandemic, no satisfactory treatment of COVID-19 was available, and the experimental off-label treatment regimens included monoclonal antibodies, antiretrovirals, and glucocorticoids [17]. While Paxlovid (nirmatrelvir/ritonavir) has emerged as a rationally developed home COVID-19 therapy for high-risk patients in early disease stages, the early approaches are still used stepwise in a hospital setting, as patients become dependent on oxygen, and as their risk for progression to severe disease increases [18]. For patients who require support oxygen, dexamethasone and remdesivir are recommended with the addition of baricitinib or tocilizumab if their requirements are rapidly increasing with signs of systemic inflammation. The next step is for patients who require high-flow nasal cannula, non-invasive ventilation, mechanical ventilation, or extracorporeal membrane oxygenation. It is recommended for such patients to receive dexamethasone plus either baricitinib or tocilizumab. For patients who are on a high-flow nasal cannula or non-invasive ventilation, remdesivir can also be an option. These therapeutic approaches are now considered standard of practice, but in the early stages of the pandemic they were not standardized, and the data on their use was not collected in a systematic fashion. The current guidelines were put in place based on several landmark clinical trials. The RECOVERY trial showed that glucocorticoids, specifically dexamethasone 6 mg daily for up to 10 days, is a preferred treatment for COVID-19. The REMED trial [19] and HIGHLOWDEXA trial [20] also supported that finding. The REMAP-CAP trial [21] showed that the IL-6 receptor antagonists tocilizumab and sarilumab decreased mortality. These agents also carry many warnings along with them, one of which importantly is increased chance of infections, especially bacterial and fungal infections. Tocilizumab carries a black box warning for an increased risk of serious infections [22].

It has now become apparent that COVID-19 is associated with the increase in systemic fungal infections, as observed by the emergence of new fungal infections such as COVID-19-associated pulmonary aspergillosis (CAPA), mucormycosis (CAM), candidiasis (CAC), as well as other infections with *Aspergillus*, *Mucorales*, and *Candida* species. We know that there are factors such as old age, diabetes, obesity, and cardiovascular disease that complicate the SARS-CoV-2 viral pathogenesis and increase the risk of severe COVID-19 infections, likely due to a compromised immune response. Risk factors for fungal infections in COVID-19 patients were shown to be old age, invasive ventilation, ventilator use, corticosteroids, tocilizumab, and broad-spectrum antibiotics [23, 24]. Several case reports and cohort studies describe COVID-19-associated fungal infections [25–29]. Most, if not all, of our current understanding of COVID-19-fungal infection association has been derived from analysis of data from multiple centers. Because the approaches to COVID-19 treatment as well as methods of diagnosis and even classification of fungal infections vary among different countries, single-center studies, where the diagnostic and treatment standards are uniform, are important to investigate relationships of COVID-19 risk factors and treatments with systemic fungal infections. Admittedly, single-center studies have an inherent drawback of a limited number of patients, reducing the statistical power. The associations of risk factors for COVID-19, COVID-19 therapies, and COVID-19 severity with systemic fungal infections have not been fully investigated. Herein, we aimed to investigate these associations and predictors for COVID-19 patients hospitalized in a US hospital.

## Methods

### Patients included in this study

This research was approved by the University of Kentucky Institutional Review Board. The data were extracted with the help of University of Kentucky Center for Clinical and Translational Science (CCTS). We considered all hospital patients who received a COVID-19 diagnosis in the University of Kentucky Healthcare (UKHC) system from April 2020 to May 2021. We considered the patients with community-acquired COVID-19. To be included, patients had to be at least 18 years old, had to have a COVID-19 diagnosis upon hospitalization and no previous diagnosis or positive test for COVID-19. Patients who did not test positive for COVID-19, those who had a previous hospitalization(s) with a COVID-19 diagnosis defined as a positive test in the previous hospitalization, those who had a hospital-acquired COVID-19 defined as a positive test 3 days or more after admission to hospital, or those with an unknown COVID-19 status were excluded from this study.

### Selection of patients with COVID-19 and systemic fungal infections

The patient data were extracted through CCTS biomedical informatics with diagnoses for COVID-19 and fungal infections identified by ICD-10-CM codes and records. A COVID-19 diagnosis was identified by ICD-10-CM code U07.1, which is specific to COVID-19, and the older coronavirus infection code B97.29 used in the initial COVID-19 diagnoses at the UKHC system. A systemic fungal infection was defined as any fungal infection, except superficial infections: *Candida* urogenital infections, *Candida* vaginal, skin, or nail infections and *Candida* stomatitis. Patients that had only superficial infections were not included in this study.

### Fungal pathogen identification

Systemic fungal infections were identified using ePlex Blood Culture Identification Panels (Roche) system. Coccidioidomycosis was diagnosed by serological testing for appropriate IgM and IgG antibodies (Meridian Bioscience). Aspergillosis was identified by a combination of imaging and sputum staining and culture tests. Histoplasmosis was identified by urine testing for Histoplasma antigen using MVista antigen kit (MiraVista Diagnostics).

### Data analysis

The data were analyzed utilizing SSPS Statistics (IBM) software version 27. Binary logistic regression was used to analyze gender and race as dependent variables and to assess predictors of a secondary fungal infection, also a binary variable. Linear regression was used for age as a dependent variable. An unpaired t-test was used to compare the continuous data between the group of patients infected with a systemic fungal infection and the non-infected group.

## Results

### Patient selection

The process of the patient selection for this study is shown in Figure 1. Between April 2020 and May 2021 there were 9,422 individual hospitalizations including two or more hospitalizations of the same patient, with 5,822 where a SARS-CoV-2 test was administered, which excluded the other 3,600 hospitalizations. Out of these 5,822 hospitalizations, 4,999 (median age: 41, F/M: 1.2/1) were included based on the criteria for community-acquired COVID-19 described in Methods. 547 patients were excluded because of a previous COVID-19 diagnosis, 241 patients were excluded for being SARS-CoV-2 negative, 34 were excluded for having tested positive ≥3 days after admission, and 1 was excluded because of the absence of test results for an administered SARS-CoV-2 test upon the hospitalization. Out of the 4,999 patients included in this study, 47 (median age: 67, M/F: 1.2/1) developed a systemic fungal infection secondary to a COVID-19 infection (0.94% incidence).

**Figure 1.**
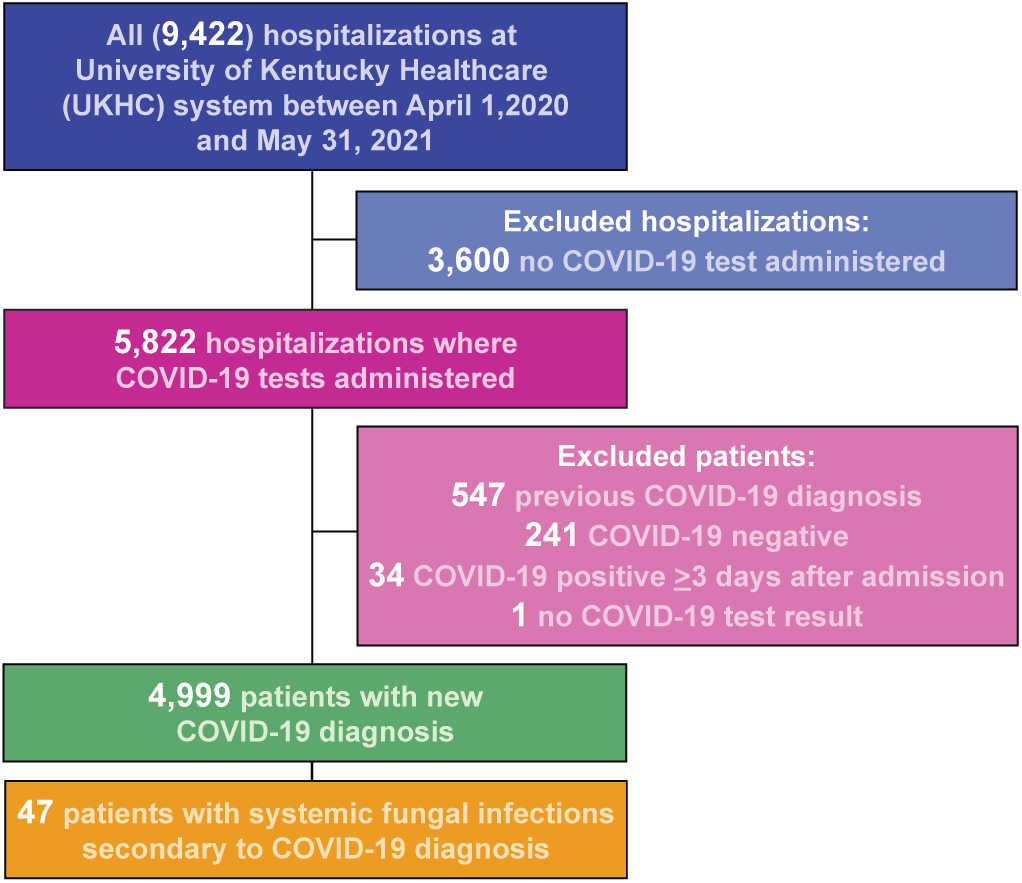
The flow chart of patient selection for this study. Out of 9,422 hospitalization records for patients ≥18 years old, 4,999 patients were included and assessed for study endpoints. 47 patients developed systemic fungal infections secondary to the COVID-19 diagnosis.

### Characteristics of patients with systemic fungal infections secondary to COVID-19

We compared the characteristics of the 4,952 COVID-19 patients who did not develop a secondary systemic fungal infection with those of the 47 patients who did (Table S1); the former group will be called non-infected and the latter infected. The gender ratios in the two groups were not significantly different. The race was self-reported; the fraction of white individuals in the non-infected group (75%) was significantly lower than that in the infected group (94%). The COVID-19 patients who did not contract a secondary fungal infection were much younger (p <0.001) than the patients who did. The age distributions in these two groups differed dramatically (Figure S1): in the very large non-infected group the maximum of the distribution was at 19 years (Figure S1A), as in the overall distribution, whereas the youngest patient in the infected group was 35 years, and the highest peak of the distribution was in at 71 years (Figure S1B). The patient heights and weights were not statistically different between the infected and non-infected groups. The proportions of those who were admitted to the ICU were drastically different, 4% and 43% for the non-infected and infected groups, respectively. The proportions of patients transferred from outside hospitals were found to be statistically significant with 12% of the non-infected patients transferred from outside hospital and 64% of infected patients transferred. The median length of stay for the non-infected group was 12 hours, significantly shorter than that for the infected group (524 hours).

### Comparison of clinical features between the infected and non-infected groups

We also analyzed the clinical characteristics of the non-infected and infected patients (Table S2). The fraction of those mechanically ventilated was 9-fold larger and the durations of mechanical ventilation were 2.3-fold longer in the group with fungal infections than in the group without fungal infections, which were significant differences. The fractions of patients with different clinical comorbidities of COVID-19 were significantly different for the non-infected and infected, with ∼4-fold larger fractions of diabetes patients, those with a cardiovascular disease and obese patients in the infected group than in the non-infected group. The mortality rates at three different time points (in hospital, 30-day post-discharge, and 90-day post-discharge) were also compared and were found to be approximately 8-fold higher for all three time points in the infected group than in the non-infected group. The OR value for mortality is approximately 12 for the infected *vs* non-infected group, making a systemic fungal infection an important predictor of mortality in patients hospitalized with COVID-19.

### The fungal infection types

The two dominant types of systemic fungal infections were candidiasis and aspergillosis. Approximately two thirds of the systemic fungal infections were candidiasis infections, while aspergillosis accounted for less than a quarter of the infections (Figure S2). Three patients were infected with two different fungal species: two with *Candida* and *Aspergillus*, and one with *Candida* and *Histoplasma*. One of these patients had both pulmonary candidiasis and pulmonary aspergillosis, and another had candidiasis sepsis and pulmonary histoplasmosis. Pulmonary aspergillosis and pulmonary candidiasis coinfections are rare, but not unprecedented in patients with severe COVID-19 [30]. As in this recently reported case, the patient died in the hospital. All aspergillosis and coccidiomycosis infections were pulmonary. Even though aspergillosis was not the dominant type of infection, it was associated with an especially high mortality risk; 4 out of 10 patients with aspergillosis died while in the hospital.

### The statistics of COVID-19 therapies and vaccinations in the non-infected and infected groups

The COVID-19 therapies administered to the patients were also analyzed for the non-infected and infected groups (Table S3). The total number of administrations of each therapy and the number of patients treated with that therapy were considered for each group. For all commonly administered therapeutics (remdesivir, dexamethasone, prednisone, methylprednisolone, and hydrocortisone) the fraction of patients that received these medications was significantly larger in the infected group than in non-infected group. Moreover, the number of administrations per patient was larger in the infected group as well.

Because there were several steroids employed, we consolidated the data on the steroid treatments by normalizing the doses of different steroids to that of dexamethasone, the currently recommended steroid for treatment of COVID-19 (Table S4). The dosing for all steroids was converted to dexamethasone equivalents utilizing the standard conversion, where 0.75 mg of dexamethasone was equivalent to 20 mg of hydrocortisone, 4 mg of methylprednisolone, 5 mg of prednisolone, and 5 mg of prednisone [31]. The conversion for fludrocortisone was based on the bioequivalence of 0.1 mg of fludrocortisone to 50 mg of prednisone [32]. The median total doses of dexamethasone equivalent were 42 mg and 95.25 mg for the non-infected and infected patients, respectively, representing a statistically significant difference between the two groups. The median for the total number of administrations, calculated using 6 mg of dexamethasone as a daily dose, was also approximately 2-fold larger for infected patients.

Finally, we examined the vaccination status of each patient and present the total number of vaccinations as well as the number of vaccinated patients (Table S5). No vaccinated patients were in the infected group, while 6.2% of the patients in the non-infected group were vaccinated.

### Predictors of systemic fungal infections in hospitalized COVID-19 patients

The above analysis shows that there is a strong association between the systemic fungal infection status and various parameters related to COVID-19 severity. We analyzed what patient characteristics and clinical indicators are predictors of fungal infections by binary logistic regression (Table S6). To aid in this analysis, we obtained OR values for the odds of a fungal infection in general as well as specifically of candidiasis and aspergillosis. Race was a weak predictor, with the OR value of ∼5, but a marginal statistical significance, and gender was not a predictor, due to a small number of patients who self-identified as non-white in the infected group. Age was a strong predictor, with ∼8 times greater odds of contracting a systemic fungal infection for a 50-years old patient or older than for a younger patient. Other COVID-19 risk factors, diabetes, cardiovascular disease, and obesity, were similarly strong individual predictors with OR values in the 6-8 range. The OR for remdesivir administration was slightly smaller. Notably, strong predictors, whose large OR values could not be explained only by their association with age were ICU admission (OR = 16), mechanical ventilation (OR = 22), and steroid use (OR = 23). ICU admission and mechanical ventilation were exceptionally strong predictors of aspergillosis (OR = 81 and 51, respectively), although, admittedly, the relatively small number of patients with aspergillosis resulted in large OR ranges. On the other hand, these two predictors were on par with age (50 and older) for candidiasis infections. The strongest statistically significant predictor was the length of hospital stay (for 13 days and longer), with the OR value of 42 for any systemic fungal infection and 32 for candidiasis. All patients that had aspergillosis stayed at the hospital 13 days or longer. We chose 13 days as a cut-off, because this was the median hospital stay for patients not infected with a fungal infection. As we noted in the previous section, the lack of COVID-19 vaccination is likely a very strong predictor of a fungal infection, but its OR values could not be calculated with high certainly because of the paucity of vaccinated patients in the infected group.

## Discussion

Several factors that increase the chances of severe COVID-19 disease have emerged from a number of clinical studies [33, 34]. Expectedly, COVID-19-associated (CA) fungal infections have similar risks among these patient characteristics and clinical features to those observed previously with influenza pneumonia [35]. Invasive pulmonary aspergillosis was a common associated infection that was initially documented in H1N1 patients [36]. It is highly important to study CA fungal infections, because such infections greatly increase mortality. Therefore, it is important to study CA fungal infection and their extent of influencing the treatment outcomes for COVID-19 patients. In this context, while multicenter studies have a benefit of large numbers of patients and robust statistics, single-center studies are useful because of the use of the uniform standards of patient care, from the admission to the hospital and the diagnosis to the discharge. In our study, the risk of death was 8-fold higher in the patients with fungal infections than in those without, reaching nearly 38%. This value is somewhat lower than the 54.6% mortality based on a review of case reports from the hospitals around the globe, but it is not significantly lower [37]. This could be explained by a higher rate of fungal infections among COVID-19 patients in Asia than in the US [38]. This study also analyzed characteristics of patients hospitalized with COVID-19 and risk factors for severe COVID-19 as well as COVID-19 therapies to examine their potential associations with systemic fungal infections. This study was a retrospective single-center study in the US, involving 4,999 hospitalized patients. The number of patients with fungal infections was significant (47, or 0.94%), allowing us to obtain quantitative OR assessment. However, even for such a relatively large study, the number of patients infected with aspergillosis was small (10), which limited the statistical power to infer some quantitative conclusions for this infection in particular. Candidiasis was by far the most common fungal infection. Age, a known risk factor for severe COVID-19, was strongly associated with secondary fungal infections. The OR for aspergillosis among older patients (50 years or older) was more than 2-fold larger than for a fungal infection in general. The strength of the associations of known COVID-19 comorbidities (diabetes, heart disease, and obesity) with fungal infections generally did not exceed that of age, even though ∼50% of the patients that had fungal infections had a heart disease and ∼50% of the patients had diabetes. Because of a strong association of age with these comorbidities, one cannot view these chronic conditions as completely independent predictors. The same was true about administration of remdesivir, an anti-COVID-19 therapeutic. Tocilizumab, an anti-inflammatory drug used in some seriously ill COVID-19 patients and known to be associated with susceptibility to fungal infection [39], was used only in 29 patients in this study, among which there were no patients with fungal infections. Four of the predictors stood out by being exceptionally strong: ICU admission, length of hospital stay, mechanical ventilation, and steroid use, for which the fungal infection OR values were higher or much higher than that for age. While all these factors could be consequences of a fungal infection added to the COVID-19 infection, some of them are likely, at least in part, causative of fungal infections. The length of hospital stay may increase chances of a transmission of a fungal infection. Ventilator tubes create a higher risk of fungus exposure and provide a surface for fungal biofilms to develop [40]. Steroids weaken immune system defenses against fungal infections. Steroid exposure was increased and the total number of doses was also higher than the recommended CDC guidelines for those that developed secondary fungal infections.(17) The median for the COVID-19 cohort was less than the total recommended dose of 6 mg once daily for 10 days, while the group that contracted fungal infections secondary to COVID-19 received higher doses. Steroids for COVID-19 are immunosuppressant and inhibit the pro-inflammatory process in COVID-19 [41, 42]. This then limits the host innate response to pathogenic fungal cells, leaving the door open for these COVID-19-associated infections. Candidiasis was a dominant type of infections, followed by aspergillosis, accounting for the majority of the fungal infections contracted by these patients. *Candida* infections have been on the rise in recent years, with *Candida auris* coming into focus due to its resistance to antifungal therapy, and its increasing incidence in COVID-19 patients [43].

## Conclusions

In summary, this study made it clear that COVID-19 patients with severe disease are at high risk of contracting fungal infections, which in turn greatly lower chances of survival for these patients. Treatment of such patients is complex and needs to carefully weigh in the risks associated with steroid use, mechanical ventilation, and a proper antifungal therapy, taking into account the fungal species and drug resistance.

## Data Availability

All data produced in the present study are available upon reasonable request to the authors.

## Declarations

### Ethics approval

This retrospective chart review study involving human participants was in accordance with the ethical standards of the institutional and national research committee and with the 1964 Helsinki Declaration and its later amendments or comparable ethical standards. The Human Investigation Committee (IRB) of University of Kentucky approved this study (IRB Number 74455).

### Authors’ contributions

All authors contributed to the writing of the manuscript and preparation of the Figures and Tables. Abbygail C. Wilbourn and Sylvie Garneau-Tsodikova conceptualized and designed the study. Abbygail C. Wilbourn and Oleg V. Tsodikov analyzed the data.

