## Supporting information for "Association of COVID-19 risk factors with systemic fungal infections in hospitalized patients"

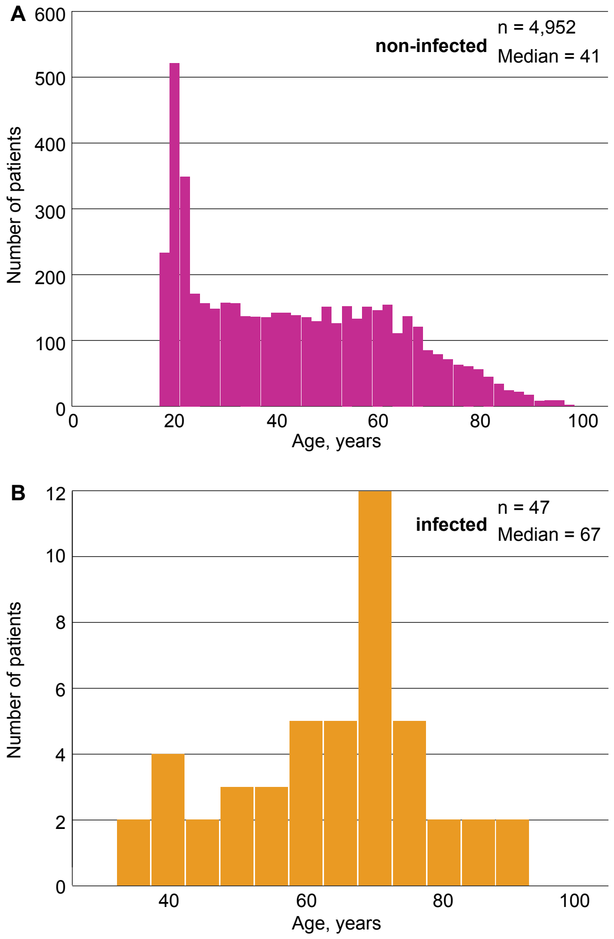


**Figure S1.** The age distribution of the patients. **A.** The age distribution in the non-infected group (pink bars). **B.** The age distribution in the infected group (orange bars).

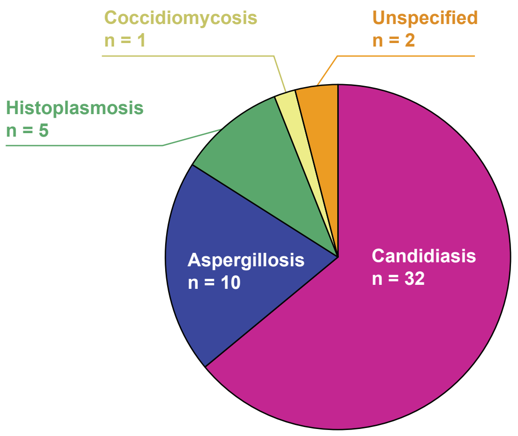


**Figure S2.** The incidence of systemic fungal infections in this study, grouped by the fungal pathogen.

### Tables

| **Table S1.** Characteristics of the patients without and with systemic fungal infections. | | |
|---|---|---|
| | **4,952 COVID-19 patients without fungal infection** | **47 COVID-19 patients with fungal infection** |
| Gender, male n (%), p = 0.194 | 2267 (46%) | 26 (55%) |
| Race, white n (%)*, p = 0.009 | 3734 (75%) | 44 (94%) |
| Median age in years (IQR)*, p <0.001 | 41 (35) | 67 (18) |
| Median height in cm (IQR), p = 0.227 | 170.0 (15.3) | 170.1 (13.4) |
| Median weight in kg (IQR), p = 0.745 | 85.2 (32.6) | 87.6 (33.7) |
| Admit to ICU, n, (%)*, p <0.001 | 222 (4%) | 20 (43%) |
| Transferred from another hospital, n (%)*, p <0.001 | 572 (12%) | 30 (64%) |
| Median length of hospital stay in h (IQR)*, p <0.001 | 12 (34) | 524 (575) |
| Abbreviations: ICU = intensive care unit, IQR = interquartile range.<br>* = statistically significant difference between the two groups | | |

| **Table S2.** Clinical characteristics of the patients without and with systemic fungal infections. | | |
|---|---|---|
| | **4,952 COVID-19 patients without fungal infection** | **47 COVID-19 patients with fungal infection** |
| Mechanically ventilated hospitalizations, n (%)*, p <0.001 | 339 (6.8%) | 29 (62%) |
| Median duration of mechanical ventilation, h (IQR)*, p < 0.001 | 208 (316) | 488 (579) |
| Diabetes mellitus, n (%)*, p <0.001 | 616 (12%) | 24 (51%) |
| Cardiovascular disease, n (%)*, p <0.001 | 684 (14%) | 26 (55%) |
| Obesity, n (%)*, p <0.001 | 337 (7%) | 14 (28%) |
| Mortality, n (%)*, p < 0.001 | | |
| In hospital*, OR = 11.0 (95% C.I.: 5.9-20.7) | 202 (4%) | 15 (32%) |
| 30-day post-discharge*, OR = 12.1 (95% C.I.: 6.5-22.2) | 222 (4.5%) | 17 (36%) |
| 90-day post-discharge*, OR = 12.3 (95% C.I.: 6.8-22.6) | 237 (4.8%) | 18 (38%) |
| * = statistically significant difference between the two groups; OR = odds ratio; C.I. = confidence interval. | | |

| **Table S3.** COVID-19 therapy administrations. | | | | |
|---|---|---|---|---|
| | **4,952 COVID-19 patients without fungal infection** | | **47 COVID-19 patients with fungal infection** | |
| **COVID therapy** | *Administrations (per patient)* | *Patients (%)* | *Administrations (per patient)* | *Patients (%)* |
| Remdesivir | 2914 (5) | 590 (12%) | 110 (6) | 18 (32%) |
| Tocilizumab | 27 (1) | 27 (0.5%) | 0 (0) | 0 (0) |
| Dexamethasone | 5472 (7) | 831 (17%) | 357 (11) | 32 (68%) |
| Prednisone | 1131 (8) | 138 (3%) | 141 (11) | 13 (28%) |
| Methylprednisolone | 2059 (14) | 153 (3%) | 213 (18) | 12 (26%) |
| Fludrocortisone | 140 (7) | 21 (0.4%) | 12 (4) | 3 (6%) |
| Hydrocortisone | 1119 (10) | 109 (2%) | 227 (18) | 13 (28%) |
| Prednisolone | 12 (6) | 2 (0.04%) | 0 (0) | 0 (0) |

| **Table S4.** Steroid administration in dexamethasone equivalents. | | |
|---|---|---|
| | **4,952 COVID-19 patients without fungal infection** | **47 COVID-19 patients with fungal infection** |
| Total mg median (IQR)*, p <0.001 | 42 (48) | 95.25 (257) |
| Max total mg | 2209 | 1081.5 |
| Min total mg | 1.5 | 7.5 |
| Total administrations median (IQR)*, p <0.001 | 7 (7) | 13 (28.25) |
| Max total administrations | 225 | 92 |
| Min total administrations | 1 | 1 |
| * = statistically significant difference. | | |

| **Table S5.** The SARS-CoV-2 vaccination status. | | | | |
|---|---|---|---|---|
| | **4,952 COVID-19 patients without fungal infection** | | **47 COVID-19 patients with fungal infection** | |
| Vaccine manufacturer | *All vaccinations* | *Patients* (%) | *All vaccinations* | *Patients* (%) |
| Pfizer-BioNTech | 409 | 308 (6.2%) | 0 | 0 |
| Moderna | 152 | 111 (2.2%) | 0 | 0 |
| Janssen | 5 | 5 (0.1%) | 0 | 0 |

**Table S6.** Predictors of systemic fungal infections in hospitalized COVID-19 patients.

| **Predictor** | **OR all infections** | **OR candidiasis** | **OR aspergillosis** |
|---|---|---|---|
| Age, ≥50/<50 | 7.9 (3.7-16.9)[a], p <0.001 | 16 (5-51), p <0.001 | 14 (2-114), p = 0.011 |
| Race, white/other | 4.8 (1.5-15.4), p = 0.009 | 4.9 (1.2-20.4), p = 0.03 | p = 0.3 |
| ICU admission | 16 (9-29), p <0.001 | 12 (6-26), p <0.001 | 81 (17-385), p <0.001 |
| Mechanical ventilation status | 22 (12-40), p <0.001 | 17 (8-34), p <0.001 | 51 (11-243), p <0.001 |
| Length of hospital stay, ≥13 days/<13 days | 42 (21-83), p <0.001 | 32 (15-70), p <0.001 | +∞;* all infected patients ≥13 d |
| Diabetes mellitus | 7.3 (4.1-13.1), p <0.001 | 7.0 (3.5-14.0), p <0.001 | 10 (3-37), p <0.001 |
| Cardiovascular disease | 7.7 (4.3-13.8), p <0.001 | 9.0 (4.4-18.4), p <0.001 | 4.0 (1.1-14.4), p <0.031 |
| Obesity | 5.8 (3.1-11.0), p <0.001 | 6.2 (2.9-13.1), p <0.001 | 8.9 (2.5-31.8), p = 0.001 |
| Remdesivir | 5.0 (2.8-9.0), p <0.001 | 4.4 (2.1-9.0), p <0.001 | 11 (3-39), p <0.001 |
| Steroids (any administration of any steroid) | 23 (10-52), p <0.001 | 14 (6-33), p <0.001 | +∞;* all infected patients + steroids |
| Lack of COVID-19 vaccination | +∞;* all infected patients | +∞;* all infected patients | +∞;* all infected patients – vaccine |

[a]The OR ranges correspond to 95% confidence intervals.
*The statistical significance and the OR range could not be calculated due to the lack of patients or only one patient in one of the categories.
